# Applying the unified models of ecology to forecast epidemics, with application to Covid-19

**DOI:** 10.1101/2020.04.07.20056754

**Authors:** Shelby Loberg, Clarence Lehman

## Abstract

During a burgeoning outbreak of a novel disease, public attention will ordinarily expand as the severity of the outbreak expands—as infections multiply and news reports accumulate. Such public attention will in turn reinforce tactics to control the outbreak. In classical epidemiological models, effects of such tactics can be incorporated in standard parameters of transmission, recovery, and mortality. Unfortunately, early in an outbreak those individual parameters may be poorly known, hence corresponding models can get lost in uncertainty. This makes it difficult to determine whether the disease is spreading exponentially or logistically, or along another path. Examining cases over time is also problematic, as a logistically growing infection that is leveling off appears exponential in early phases. Here we report on the most basic mechanistic, ecological model we can imagine, which can help distinguish growth that is and is not under control. This approach did a satisfactory job predicting the final outcome of the Ebola outbreak of 2014–15. The model’s two parameters were computable in real time, well before the outcome was actually known. The first parameter is an intrinsic rate of increase in cumulative deaths or reported cases. The second parameter is related to the human social system and represents all tactics that combine to control the outbreak. That parameter is coupled to the number of cumulative deaths or cases. We examine the basic mechanisms operating in this model and show the predictions made during the Ebola outbreak. We also consider how this basic model is performing for the Covid-19 pandemic and highlight ecological models that align with popularly discussed concepts such as flatten-the-curve, exponential growth, and inflection points of curves.

**Highlights:**

- We exhibit a macroscale model that includes a biological pathogen-growth factor and an ecological social-response factor.
- All parameters in the model are designed to be measurable from publicly available data.
- Early in the 2014–15 Ebola outbreak the model accurately predicted the final number of deaths.
- The same model displays a low chance of 100,000 deaths or more in the United States if strong measures continue and expand.

## I. Introduction

In September 2014, some Ebola modelers were allowing for worst-case scenarios, partly by projecting continuing exponential growth [18], and later received discouraging coverage for overestimating the outbreak [6]. The number of cases of Covid-19 are often considered exponential as well, and this has major repercussions, for example, in communication to the public, projecting morbidity and mortality, and evaluating effectiveness of control measures. However, the early phase of logistic growth also appears exponential. How can the two be distinguished? Rather than determining when the exponential part of a disease curve will “switch” to logistic, we propose considering the more likely scenario that disease cases and other single populations in nature never grow exponentially—that is without bound or carrying capacity—for long [7]. Although there is an early phase of logistic growth that appears “exponential-like” (and some populations may actually grow exponentially or super-exponentially at times) the growth of an epidemic is likely logistic throughout. The basic method presented here revealed that the spread of Ebola in 2014–15 was logistic from early in the outbreak. That is, the rate of deaths were declining relative to new deaths from the beginning, undoubtedly due to ever-increasing efforts at control. From early in the outbreak, the method projected final deaths in the epicenter of western Africa to be the order of 10,000 (Figure 1C), which is close to how the outbreak developed. What will the same method say about Covid-19?

**Figure 1:**
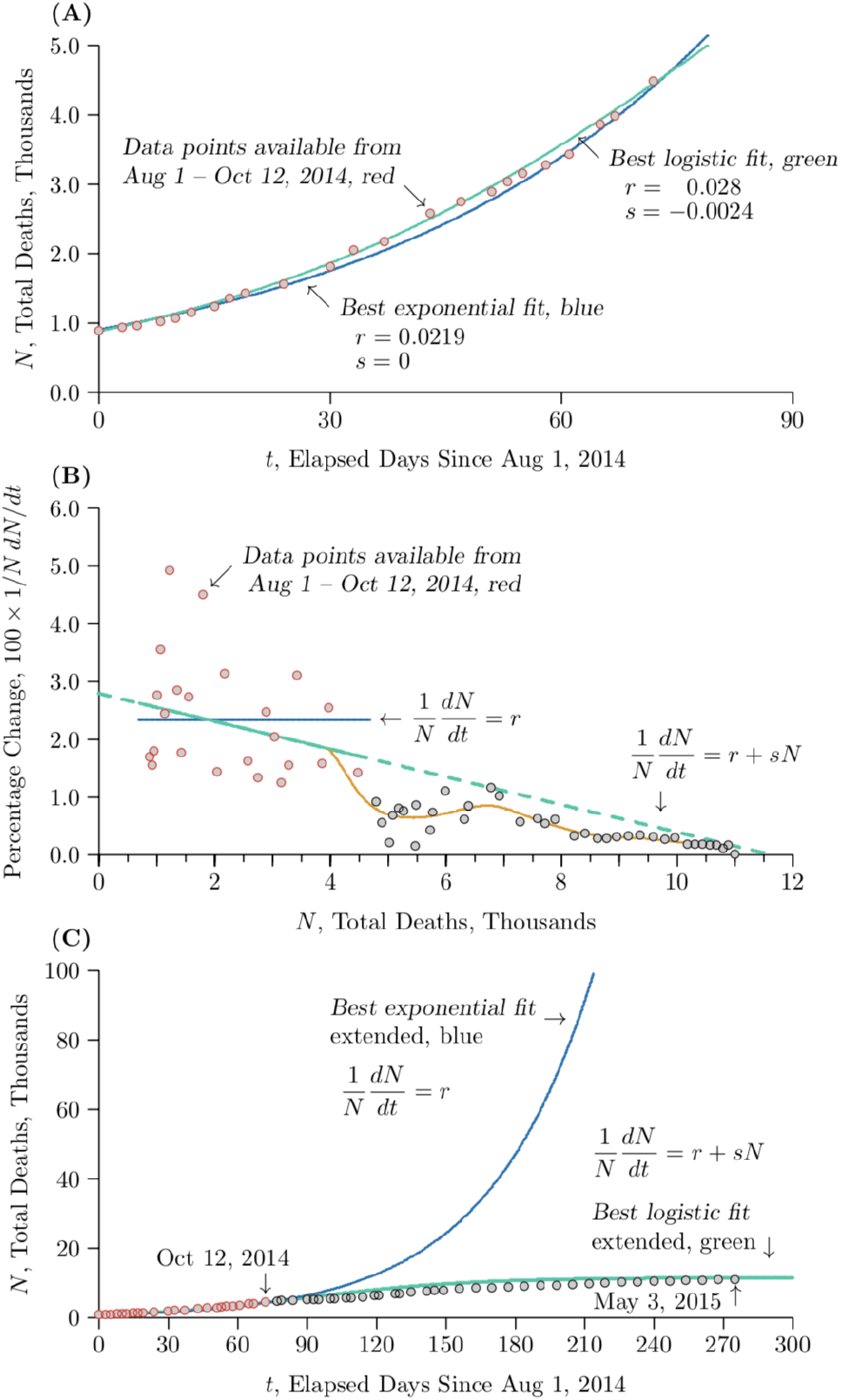
Early projections of the Ebola outbreak of 2014–2015. Red and gray dots mark empirical estimates of cumulative deaths from who sources as the outbreak proceeded. The red dots were used to generate the projections, the gray dots to judge accuracy. Solid curves show various projections. **(A)** Deaths were accelerating when we started tracking the outbreak as part of a classroom exercise, though at a decelerating relative rate. The blue curve is the best-fit projection based on doubling times among the red dots—in other words, estimating the exponential growth. The green curve is the best fit for logistic growth. The blue and green curves seem quite similar, but they have dramatically different outcomes rather quickly. **(B)** The same data shown as percentage change in deaths versus total deaths. The red dots from early in the epidemic reveal a downward slant. The green line is a regression line that follows this trend, whereas the blue line is the mean through the red points. Using the mean generates exponential growth, and using the slope generates logistic growth. Despite much noise in the early data, the green regression line successfully projected the outcome. Looking at the data points, a downward blip began between four and five thousand deaths, and then an upward super-exponential blip arose, followed by another downward and smaller upward blip before settling down (traced in orange as a curve smoothed from the data). Curiously, this roughly matches the theoretical pattern illustrated in Appendix II. **(C)** Early projections and the actual course of the outbreak. Blue is an exponential projection from the red dots at the beginning of the outbreak, corresponding roughly to other early estimates of an emerging pandemic. This is the same curve as in Part A but extended forward in time. Green is the logistic projection from the rsN model on Oct 12, 2014, again based only on the red dots. Gray dots show the actual course of the outbreak, in reasonable correspondence with the theoretical projection in green.

## II Models

A model is just a simplified view of something more complex. One of the early disease models was by William Farr, who collected data on smallpox deaths in the 19th century [17] and found the time course had the shape of a Gaussian curve. That became known as Farr’s Law and is still referenced today [20]. It is entirely phenomenological, meaning that it simplifies the description of the phenomenon, rather than proposing a simplified mechanism underlying the phenomenon, as a mechanistic model does.

Ideally, a mechanistic model provides (1) a simplified view of a complex system, and (2) glimpses into the future of the system based on measurements from the past and present. To accomplish the second point, a mechanistic model must have dynamical variables and parameters that can be measured. If it is overly mechanistic—if it contains elements that cannot be readily determined early in the outbreak of a novel disease—it may help understand the conceptual dynamics of a system, but cannot be used for projections of the system into the future. Even basic epidemiological models, such as the common *SIR* model (Appendix I), are overly mechanistic in that way. Of course, for a conceptual understanding of disease transmission, this representation is used because new infections in susceptible individuals are caused by individuals previously infected, so the development of the disease depends on both of those numbers. However, without medical tests able to detect infections, the number infected will not be known, hence the transmission parameters such as rates of infection, recovery, and death will not be known. Thus it can be difficult or impossible to accurately project the course of the outbreak using a classical model with multiple unknown parameters [9]. In addition, early in the epidemic, individual variation in transmission and behavior may have outsized effects, making them even more difficult to parameterize. Therefore, simpler views are needed.

Here we apply a simpler view based on the unified models of ecology [15]. In their most basic form applied to epidemiology, these contain (1) a biological parameter, *r*, related to the infectivity of the pathogen, and (2) an ecological parameter, *s*, related to interactions among the hosts. Whether the hosts are crops or other plants, wild or domestic animals, or humans, the ecological factor is essentially an interactive or social factor, which becomes most prominent in human responses to serious diseases.

In this paper we are using macroscale models, which amalgamate all the individuals of a category into a single number representing that category, such as the total number of deaths at a given time. In contrast, microscale models [10], such as agent-based models, keep track of individuals, thus allowing variations among individuals. Microscale models can handle genetic variation within groups, variation in infectivity among individuals, such as super-spreaders in disease, and other finer details. A macroscale model is often a few lines of computer code (Figure 5), whereas a microscale model can be many thousands of lines of code, and therefore not always helpful early in an epidemic when many details remain unknown.

However, a macroscale model will not tell how many hospital beds are needed in specific locations, in which specific regions within a country outbreaks will occur first, or what specific measures should be taken to combat a disease. What it can tell is the magnitude and timing of the problem, to help inform all the conflicting requirements to combat an expanding disease while keeping a society functioning. It also tells if a complex microscale model is in the right ballpark. To be believed, we propose that a complex microscale model should be paired with a corresponding transparent, empirically based macroscale model, whose output the microscale model matches when its internal conditions are sufficiently simplified.

With respect to Ebola in 2014, the number of infections early in the outbreak was not accurately known. But deaths occurred in a fraction of those infected and could be tracked as a more definitive measure. When properly rescaled, the cumulative number of deaths can be a proxy in particular models for the primary dynamical variable, the number of infections (Appendix I). Moreover, in a dread disease like Ebola, the cumulative number of deaths affects society’s view of the seriousness of the outbreak, and hence the number of deaths feeds back into efforts to control the disease. Cumulative deaths thus becomes a primary dynamical variable in its own right, and secondarily a proxy for the number of infections at a fixed average time in the past.

How can cumulative deaths be a variable in a dynamical system, when prior deaths do not directly cause new deaths? This is because at the early stage of an epidemic when the growth rate appears approximately exponential, the cumulative number of deaths is mathematically the integral of the current number of deaths, and for the exponential function those two are proportional (Appendix I).

We create our macroscale model as a simplified *SIR* model. Early in the course of a rapidly spreading outbreak of infectious disease, the fraction of the population infected is low, which simplifies the dynamics and reduces the number of parameters needed to estimate the ultimate size and duration of the outbreak, and thereby help determine the level of human effort needed to control it. In such situations the standard *SIR* model can be greatly reduced to a related but rather different “*rsN*” model, which is amenable to modeling and data fitting, even early in the course of an outbreak, because its parameters can be known. It is the simplest form of a density-dependent ecological model, motivated by a general model for growth of an ecological population as a function of its density (Equation 1 of Figure 2, [12]). Truncating on the right of the equal sign to terms of first order creates the two-parameter *rsN* model (Equation 2 of Figure 2). With its two-parameter structure, its differential form has an explicit solution as a function of time, shown as Equation 3 of Figure 2.

**Figure 2:**
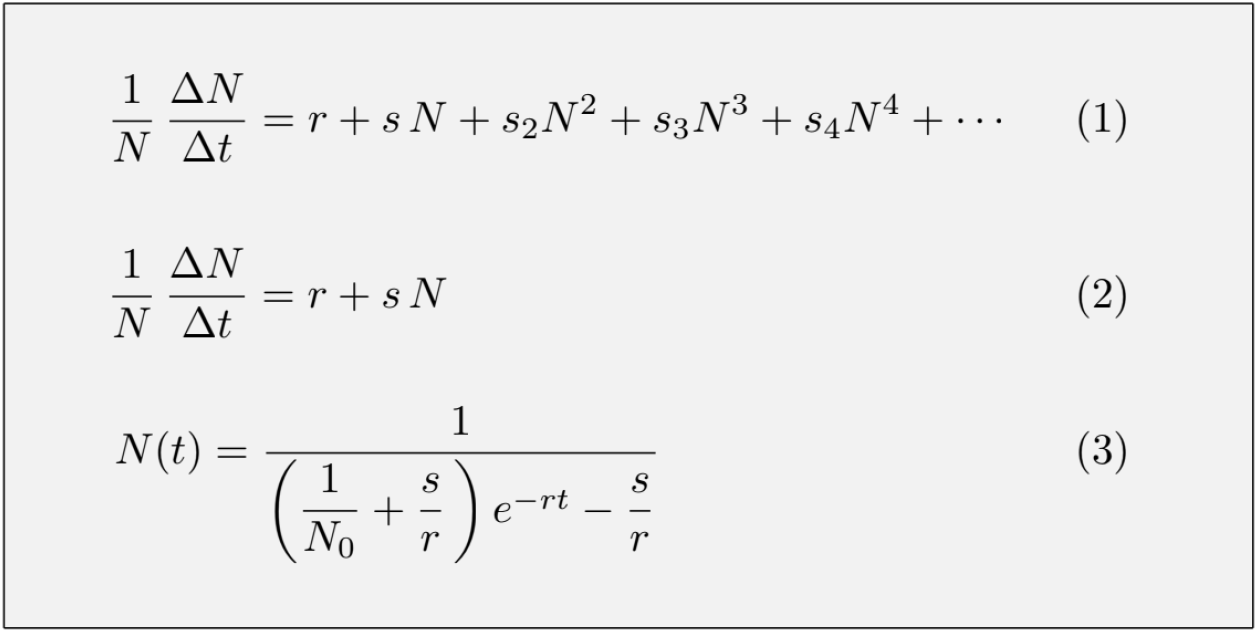
One-dimensional unified equations of ecology. Equation 1: General polynomial form for any one-dimensional ecological dynamics. Equation 2: Basic rsN form applied here. Equation 3: Explicit solution to the differential form of Equation 2. Technically, ΔN and Δt apply to finite intervals that arise in actual data, whereas dN and dt refer to the differential equation models’ smoothing of them. For the purposes of this paper the two notations can be considered interchangeable.

The term to the left of the equal sign in Equation 2 of Figure 2 represents the relative growth in the number of individuals included in variable *N*. The two terms to the right of the equal sign in the same equation, *r* + *sN*, represent an exponential growth rate, *r*, plus a modification to that rate, *sN*, varying as the density *N* varies. We consider *s* to be an ecological interaction parameter, related to the effect of the pathogen and the human response to the pathogen. We consider *r* to be a biological parameter, related to the intrinsic properties of the disease and its spread when conditions are ideal for it ([14], [15]).

Under the conditions that prevail at an early time *t, N* is small and all the terms involving *N* on the right of Equations 1 and 2 of Figure 2 are insignificant, leaving the intrinsic rate *r* as the dominant parameter. Likewise, if the *s* term is zero, density has no effect at all. In those cases Equations 2 and 3 represent exponential growth, with a fixed doubling time ln 2/*r* that does not change as the outbreak proceeds.

If *s* is negative, then Equations 2 and 3 of Figure 2 represent logistic growth, which on an *N* versus *t* graph approaches a horizontal asymptote that is called the carrying capacity. The equivalent in epidemiology would be prevalence of an endemic infection if *N* records cases, or no further deaths from the disease if *N* records deaths. On the other hand, if *s* is positive, then Equation 2 represents what is called “orthologistic” growth, so named because on an *N* versus *t* graph it approaches a vertical asymptote, orthogonal to that of the logistic equation, indicating a time before which the equation no longer applies and a different equation comes into play ([15]).

We should note that in ecological circles the logistic equation is sometimes thought of as non-mechanistic. That comes in part because it usually has not been written in per-capita form, but in a quadratic form to the right of the equal sign, which does not separate the biological from the social components.

For data in the Ebola outbreak, we used time points and number of deaths, but cases could also be used if accurate data were available. We then calculate (1) intervals of time, Δ*t*, between successive observations, (2) the cumulative number of deaths, *N*(*t*), at each time, and (3) the increment in number of deaths for each period, Δ*N* (Appendix III).

We plotted the results on a graph where the vertical axis shows the relative growth rate—that is, the per-capita growth rate 1/*N* Δ*N*/Δ*t*. The horizontal axis shows deaths (or cases) thus far, *N* (Figure 3E, F). Because Equation 2 of Figure 2 is linear to the right of the equals sign, we then fitted a line to those data. This line has three valuable properties. First and most important, its horizontal intercept estimates the number of deaths that will have occurred, if the fitted parameters reasonably represent reality, when the outbreak would finally be subdued. Before the time of that intercept, expanding efforts to control the disease will have driven the effective value of the basic reproductive number, *R*_0_ [1], below 1. Second, its intercept with the vertical axis shows the intrinsic growth rate *r*—the rate of increase in deaths relative to the number of deaths. Finally, its slope represents the strength of the social response, *s*. On such axes, exponential growth (*r >* 0, *s* = 0) appears as a horizontal curve (Figure 3F), logistic growth (*r >* 0, *s <* 0) appears as a downward sloping curve (Figure 3E), and orthologistic growth (*s >* 0) appears as an upward sloping curve (not shown). Orthologistic growth can appear in microbes or other organisms that have a mutualistic relationship with the host [4]. Thus plots on such axes distinguish among three important classes of ecological dynamics, even when the differences are subtle and cannot be readily perceived in most other ways of examining the data, such as trying to track doubling times.

**Figure 3:**
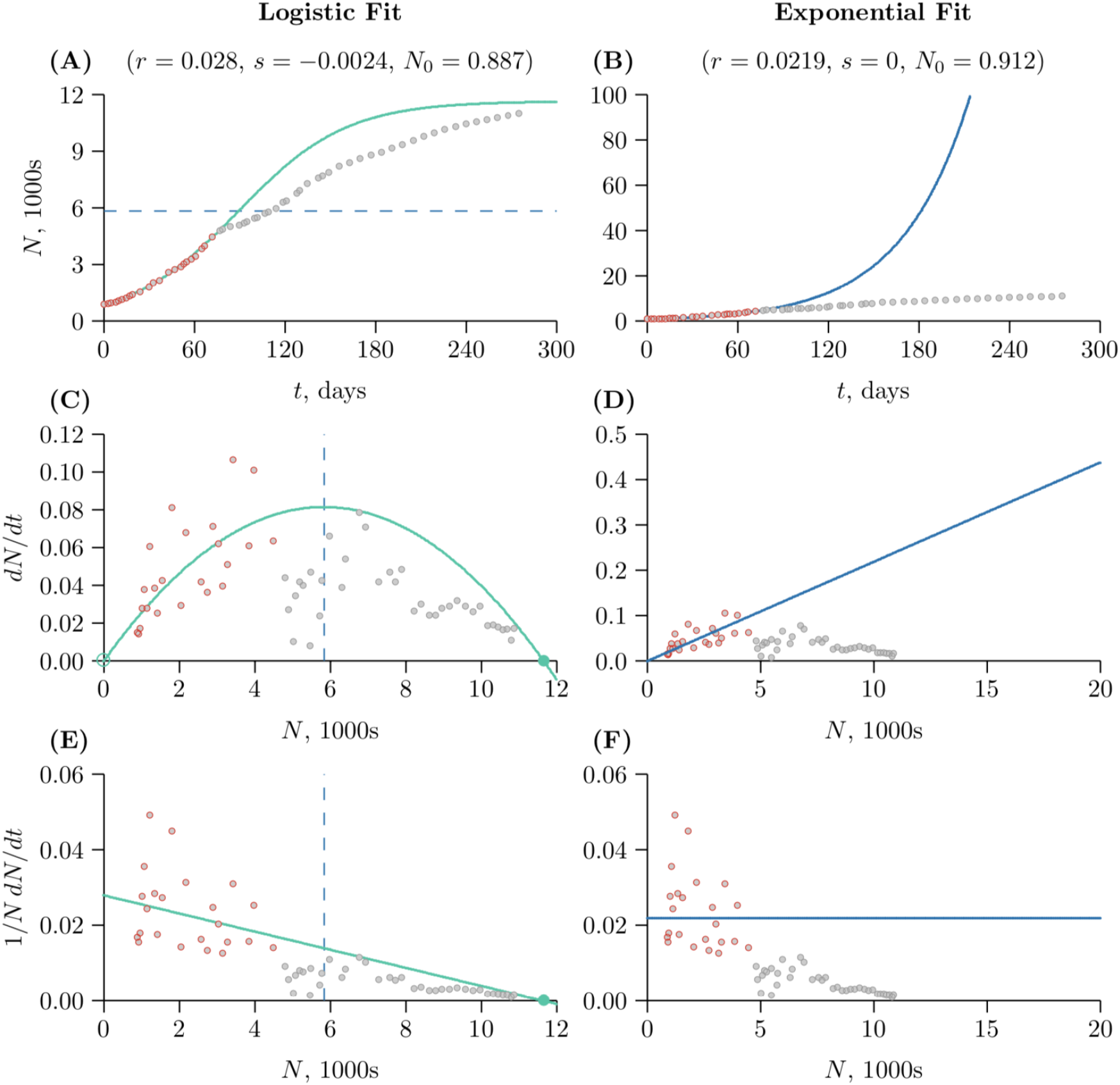
Comparing the fit of logistic and exponential growth for Ebola 2014–15, viewed three ways. The left column illustrates logistic growth, and the right column illustrates exponential growth. The red dots were used to generate the projections, the gray dots to judge accuracy. The first row is population size N versus time t, the second row is population growth rate ΔN/Δt versus population size N. The third row is per-capita growth rate 1/N ΔN/Δt versus population size N.

As a closing point on models, it is interesting that Farr’s law, which is entirely phenomenological, can be recast into a mechanistic form. In the present Covid-19 pandemic, fitting cumulative Gaussian curves to the data for cumulative deaths [19] parallels what Farr did for monthly deaths. However, a cumulative Gaussian curve is essentially indistinguishable numerically from a properly parameterized logistic curve [3], and therefore what Farr did, and what has been done subsequently, can be considered to be fitting the *r* and *s* parameters of a mechanistic ecological model, using Gaussian curves as surrogates. The *µ* and *σ* of the cumulative Gaussian curve map over to *r* and *s* of the logistic curve, and hence in this sense Farr’s method has been mechanistic all along.

## III. Lessons from Ebola

The first step we followed to project the Ebola outbreak of 2014–15, as an ongoing exercise in classes on ecology of infectious disease, was to plot the cumulative number of deaths as a function of time. Figure 1A shows that curve as it appeared in October 2014. It appears to be exponential, and indeed an exponential curve (blue) fits with *R*^2^ *>* 0.99. However, we expected that if the number of deaths *N* would continue to increase, more and more international effort would be placed on combating the outbreak. Therefore, the social parameter, *s*, should be negative, even though the value or even the presence of *s* is not clear from the cumulative curve of Figure 1A. We included no values before August, 2014, because some of those values were reportedly computed and we did not want to mix two models together—the model to compute the early estimates and the model we were creating. We started with about ten weeks of data to project the remainder of the outbreak.

When the resulting per-capita values, 1/*N* Δ*N*/Δ*t*, were plotted against deaths, *N*, as in Figure 1B, amidst all the noise, a trend appeared. The first projection we obtained in class, in October 2014, was the order of 10,000 deaths (intersection of the solid green line in Figure 1B with the horizontal axis). Because of the considerable noise in the early data, the projection was quite uncertain. However, the probability that it would be an order of magnitude or more higher than 10,000 deaths seemed low.

The next step was to take the estimates of *r* and *s* obtained by the process of Figure 1B and use them as starting points for a nonlinear fit of Equation 3 of Figure 2, the solution to the differential form of Equation 2. That results in a slight refinement of the estimates of *r* and *s*. We then used Equation 3 to project and track the outbreak as time passed. Figure 1C shows that result. The blue curve shows an exponential increase in deaths based on fitting a horizontal line through three months of points in Figure 1B. The green curve shows that the projection we made in October 2014, assuming that the parameter *s* had meaning in representing an ongoing response to the outbreak. The red dots represent the data we use to make both of our projections, and the grey dots are the actual data available as the outbreak ran its course.

For a few weeks the actual data could not distinguish the curves, but within a month it was becoming clear that there could be merit in the more modest projections of logistic growth. This was, by the way, a source of considerable motivation for students in the class, who began to realize they would be able to project such a complex thing as the course of a disease outbreak using the basic ecological tools they had been learning, as well as a source of cautious hope that a global pandemic might not actually develop for Ebola, due to the increasing efforts we could tell were happening in the world by looking at the steadily negative value of *s*.

## IV. Application to Covid-19

We are now following the same steps outlined above to model Covid-19. Just as with Ebola, we chose to use cumulative deaths available daily by country for this model [13], [8], [19] although the CDC acknowledges that deaths from Covid-19 may be underestimated [5]. We found theoretically that including discrete regions or countries can result in temporary periods of orthologistic growth moderated by subsequent periods of logistic growth (Appendix II). To model that calls for a metapopulation model [16], or possibly a spatially explicit model with a fat-tailed distribution for dispersal, but there would not be enough information to parameterize such a model at the beginning of the spread, and indeed some of the important dispersal events are intrinsically stochastic. But as shown in Appendix II, a disease spreading sub-exponentially within each region, and spreading sub-exponentially from region to region, can leave a signature of periodic super-exponential growth across all regions, even as the disease is moderating in all regions. Indeed, such a signature appears to be present in the Ebola data (Figure 1B).

China is an illustrative case because of its mature epidemic. The slope, *s*, of a regression line fitted to 1/*N* Δ*N*/Δ*t* versus *N* was negative, indicating logistic growth, since at least early February (Figure 7). For example, a regression line through points on days 25 (mid-February) to the present projects final deaths to be above 3000. This can also be calculated by *− r*/*s*. Other countries, for example Spain and Italy, also have a negative *s* term, indicating that the epidemic may be coming under control even as deaths accumulate (Figure 7). However their curves recently appear to be turning horizontal, indicating a departure from the logistic curve and corresponding control of the epidemic.

Assuming the signature of logistic growth for the US epidemic is meaningful, the regression line through daily per-capita death rates between late March and mid April in the US projects a range of potential deaths. The choice of data points to include in the regression line is as yet unsystematic, so it is more useful to examine ranges of final deaths. Figure 4 shows linear regressions run through random subsets of the data points shown as dots. A histogram for final deaths in the US is shown in Figure 4.

**Figure 4:**
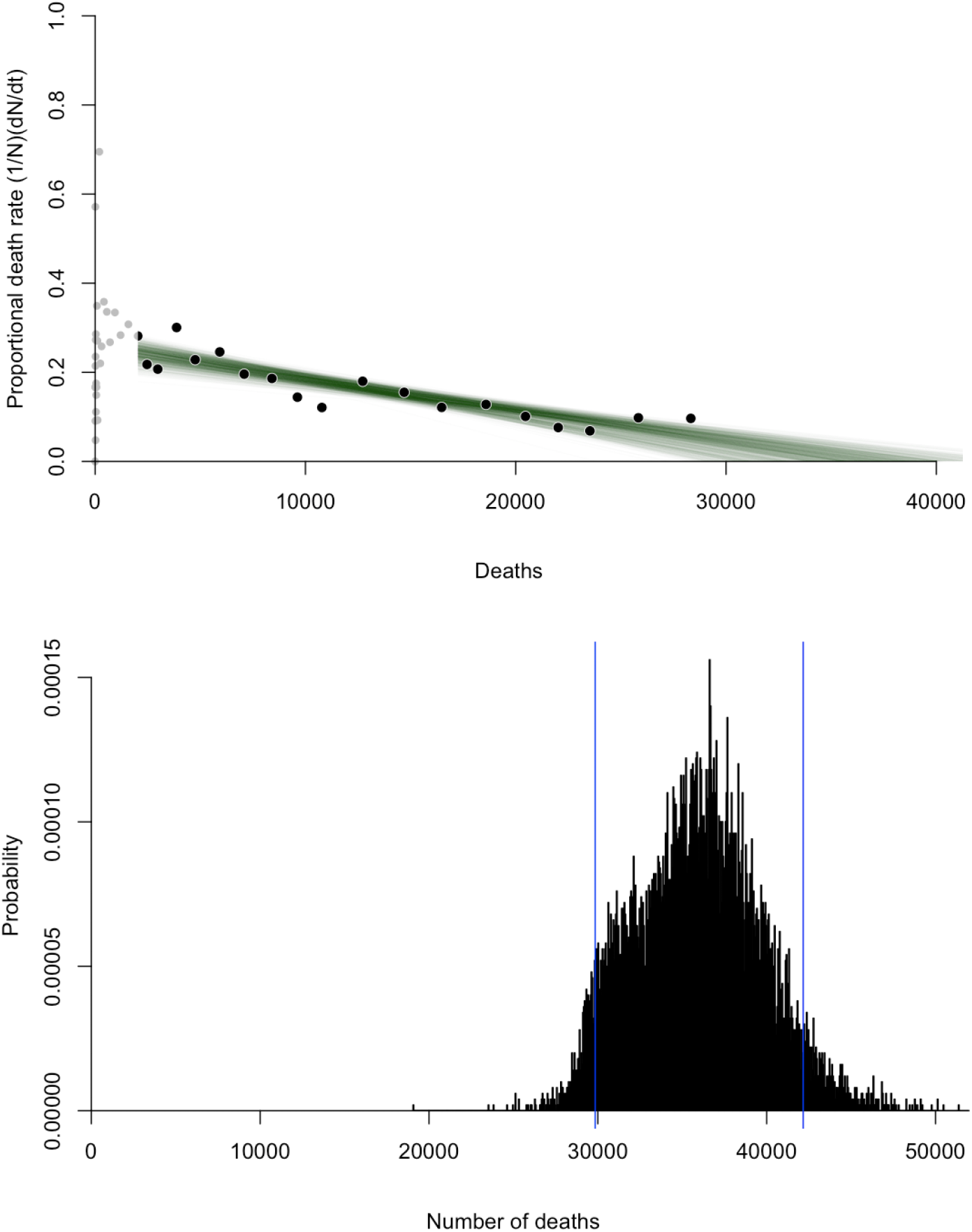
(Top) Projections of total deaths from Covid-19 in the US. The intercept of the green regression lines with the horizontal axis is a possible outcome for the US, but the waves in the data points above and below the regression line suggest the outcome is uncertain. The regression lines were drawn through a random subset of the filled dots. (Bottom) Histogram of the projected outcomes in the figure above. The figures were made from an application we created, publicly available at z.umn.edu/covid-19-rsn to examine up-to-date versions of the graphs shown here, which are necessarily frozen at the time of this writing.

**Figure 5:**
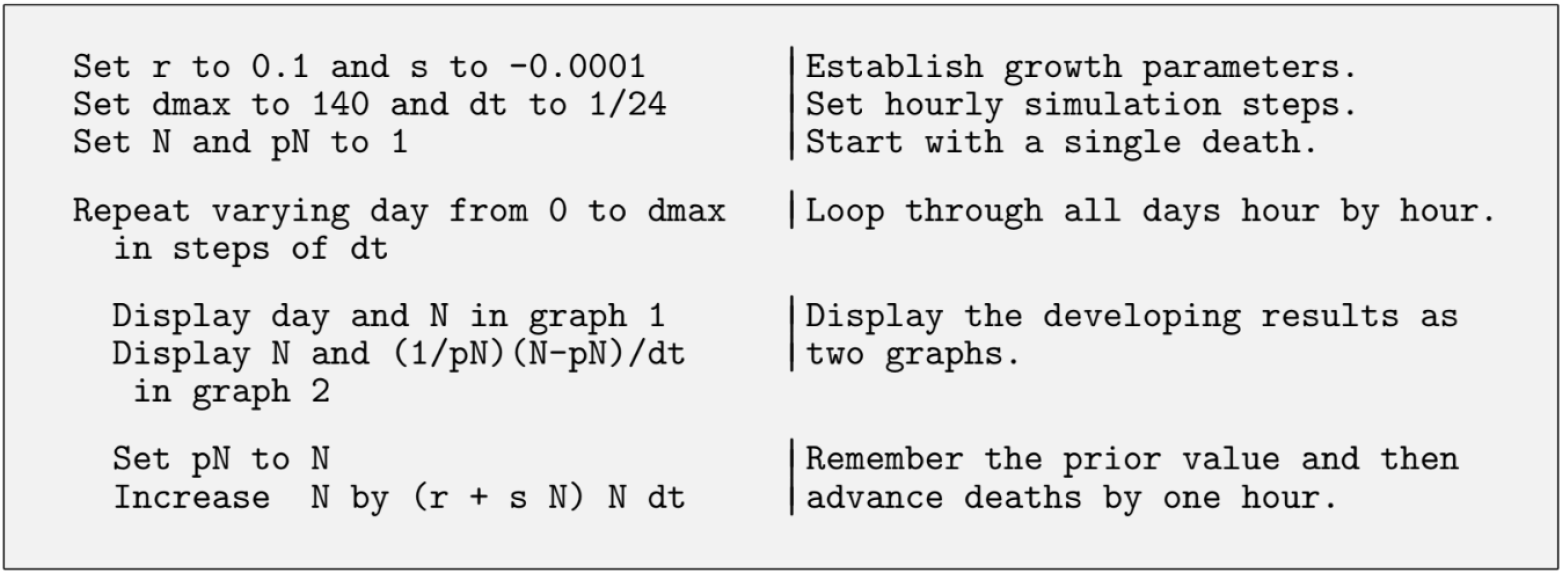
Simulating an rsN model. With r of 0.1 and s of − 0.0001, the simulated outbreak stabilizes at 1000 deaths, equal to − r/s. This generic pseudocode uses the Euler method to create two graphs, one showing the number of deaths as a function of time, due to the r term, and a second showing a steady decrease in the relative rate of deaths, due to social efforts advancing to combat the disease, encoded in the sN term. There are more sophisticated methods than Euler’s, but with the present speed of computers, and with small enough time steps, the basic Euler’s method suffices for most needs.

## V. Signatures of growth

After extracting estimates of *r* and *s* from regression lines shown in Figure 7, the parameters can be used in other ecological population growth models. Figure 8 shows three examinations of population growth comparing China and the US. Panels (A) and (B) are cumulative deaths, *N*, over time in days. Days begin from January 22 ([13], [8]). Sample simulation code appears in Figure 5.

**Figure 6:**
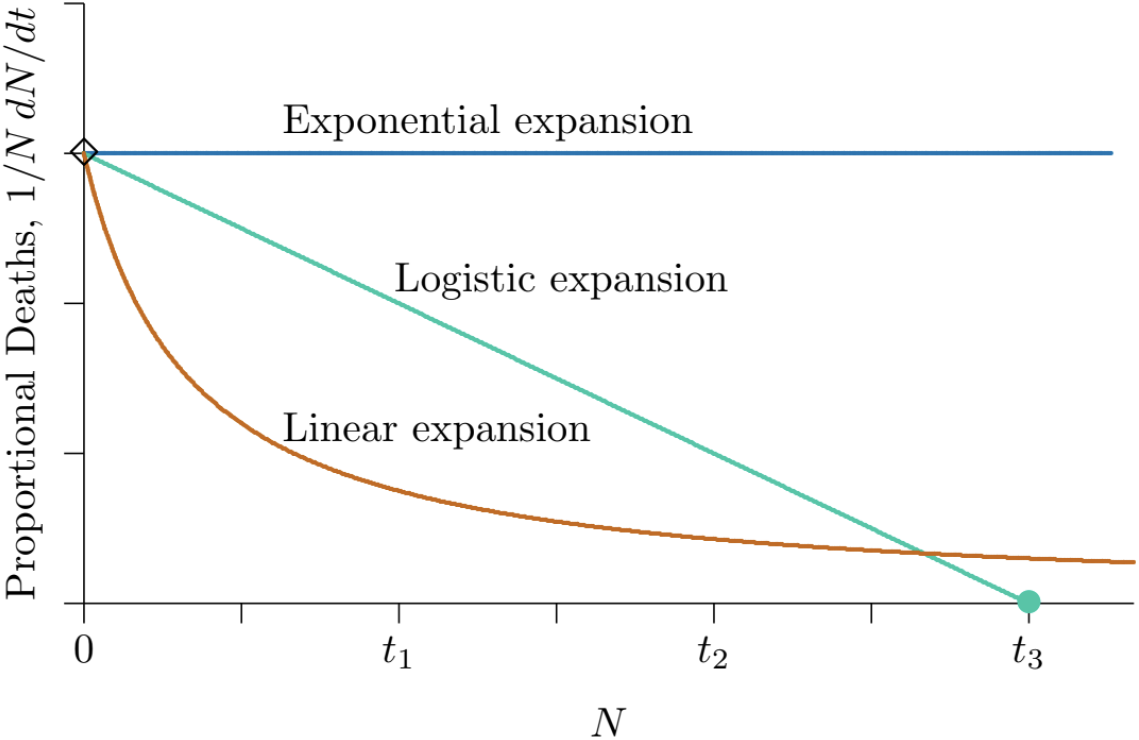
Signatures of growth for possible control measures. Exponential expansion of an epidemic can result from no special efforts to control a disease (blue line), linear expansion can result from fixed efforts (red curve), whereas ever-increasing efforts are called for to extinguish the disease (green line, touching the horizontal axis in a finite time).

**Figure 7:**
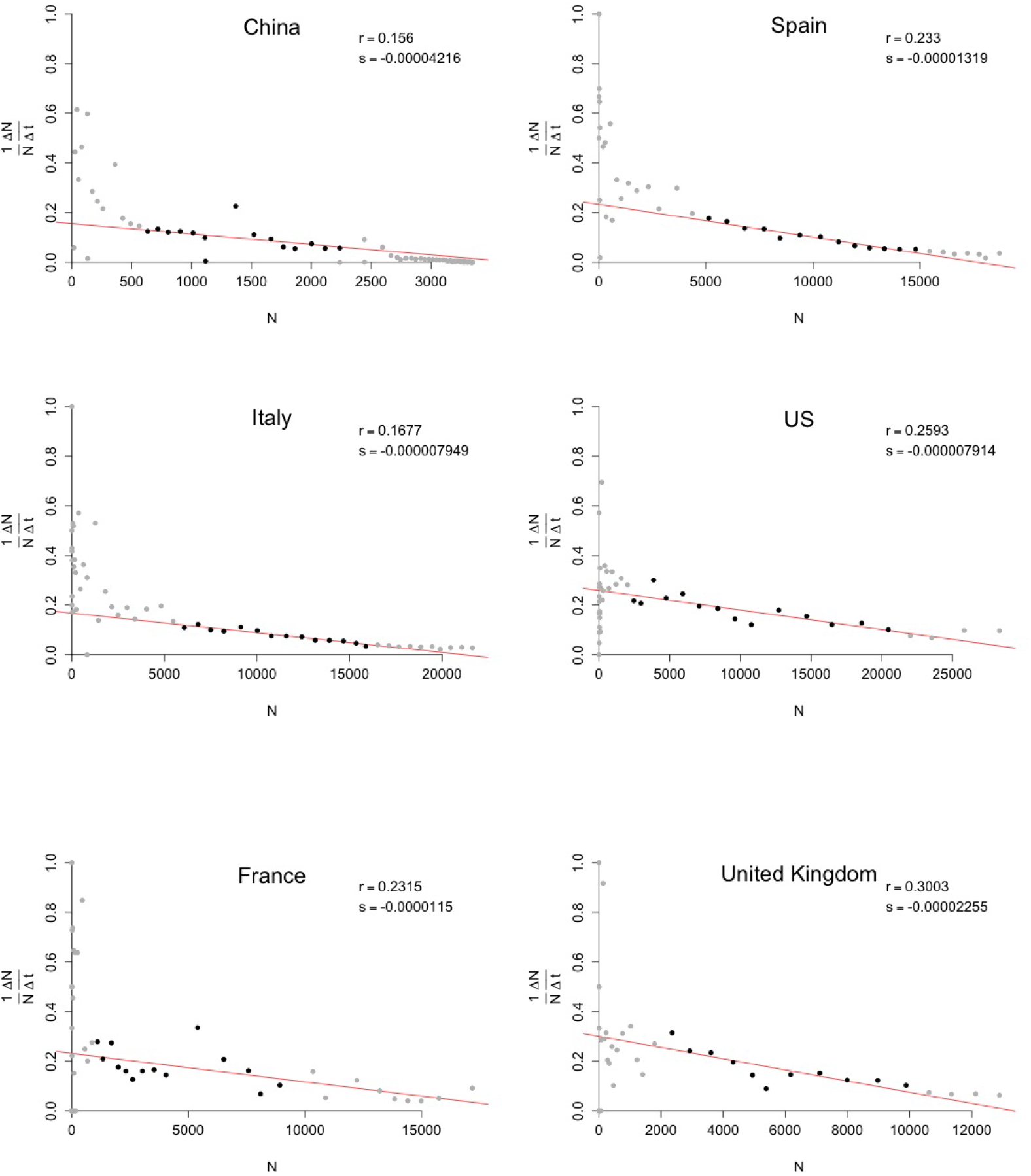
Per-capita death rate 1/N ΔN/Δt versus deaths N reported from Covid-19 in some countries with the highest death tolls as of the date of this writing. Day 1 is January 22, 2020, in each panel. One sample regression line indicates a possible trend for each country. A horizontal regression line indicates exponential-like growth of the disease and a negative slope indicates logistic growth. Data above the curve as it approaches the axis indicate growth departing from logistic.

**Figure 8:**
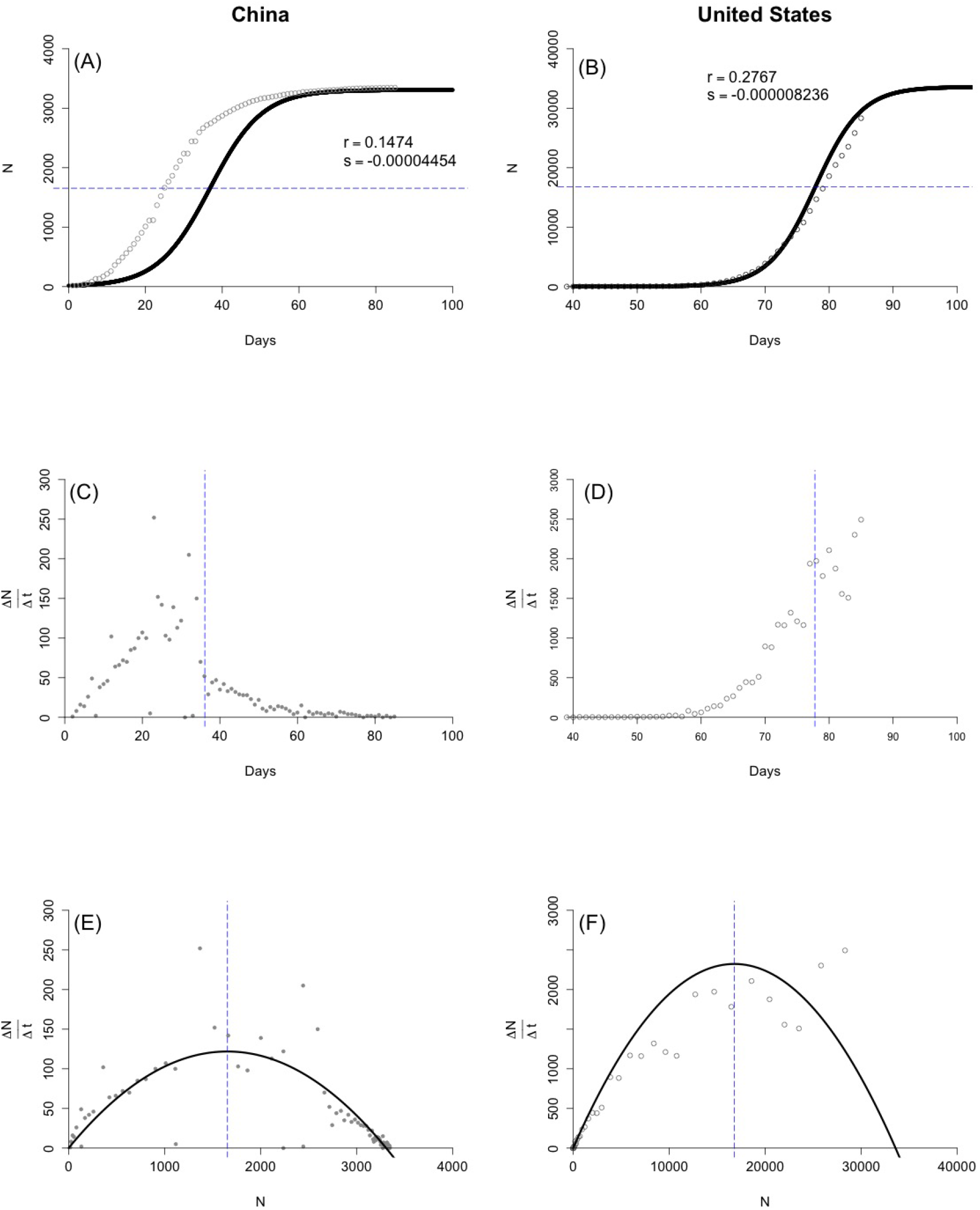
A progression of plots comparing Covid-19 deaths in US and China. Panels (A) and (B) depict deaths, N, over time in days. Day 1 is January 22, 2020 in each panel. A horizontal blue line indicates one possible inflection point where ΔN/Δt is at a theoretical maximum, which is at half of projected equilibrium N. Panels (C) and (D) depict population death rates, ΔN/Δt, versus time. The blue line depicts the theoretical time at which population death rates, ΔN/Δt, are at maximum using a likely r and s value. In the time following the blue line, deaths per day are projected to decrease, indicating a “flattening of the curve.” Panels (E) and (F) depict population death rates, ΔN/Δt, versus deaths, N. The blue line indicates the same maximum ΔN/Δt.

Panels (E) and (F) of Figure 8 depict population growth rate, Δ*N*/Δ*t*, versus population size, *N*. That is, the number of reported deaths per day versus the cumulative number of deaths. The theoretical logistic growth plotted on these axes is a second-order curve with horizontal intercepts where new deaths per day is zero. The blue line is the inflection point of the curve and corresponds with the horizontal line in panels (A) and (B).

At the inflection point of a logistic curve (see also blue lines in Figure 3A, C, E), the population growth rate Δ*N*/Δ*t* is at its peak. That is, the number of new cases (or deaths) per unit time is at its maximum. In the theoretical form, the inflection point corresponds to half the carrying capacity. The inflection point does not indicate a change in growth pattern, say from exponential (Figure 3B) to logistic growth (Figure 3A). Instead, in this representation the epidemic growth has been logistic all along, so the approximate location of the inflection point can be estimated early on, assuming consistent conditions. There are two steps to calculating the theoretical peak population growth rate, Δ*N*/Δ*t*, or the inflection point of the logistic curve. Find the carrying capacity, *K*, where the regression line in the 1/*N* Δ*N*/Δ*t* versus *N* graph crosses the horizontal axis. This can also be calculated as *−r*/*s*. Next divide that value for *K* in half, *−*(*r*/*s*)/2.

Panels (C) and (D) of Figure 8 depict population growth rate, Δ*N*/Δ*t*, versus time in days. This is the number of reported deaths per day over time. The familiar phrase “flattening the curve” applies to this graph. The blue line is analogous to the blue lines in the other panels of Figure 8; it is the time where Δ*N*/Δ*t* is theoretically at a maximum. After the time indicated by the blue line, if the *r* and *s* are accurate, the daily number of deaths will decrease.

## VI. Overcoming an epidemic

William Foege, who helped orchestrate the complete elimination of smallpox from human populations in the world, explains that we can conquer disease because we evolve more rapidly than the pathogens do [11]. That may seem surprising and impossible, but he points out that pathogens are restricted to adapt biologically, whereas we can adapt socially and technologically much more rapidly than that. Thus we overcome.

In order to overcome and eliminate a deadly disease from a region in a finite time, the proportional rate of deaths for that region must reach zero in a finite time, as shown globally for Ebola-14 in Figure 1, or as China has approached for Covid-19 in Figure 7, or in the green theoretical curve of Figure 6. Many countries appear to be on such paths now, but some instead show straight-line increases in cumulative number of deaths when plotted against time [2]. A straight-line increase in cumulative number of deaths, or equivalently cumulative number of cases, indicates that control measures are in place in the country, but that those control measures may not be sufficiently advancing with time. The corresponding curve has a hyperbolic shape (red curve in Figure 6). If that continues, the disease may become endemic, as so many diseases have become throughout the course of history—embedded in the population at some average prevalence, often with periodic outbreaks.

The message here is that for a disease to be conquered, efforts to control it must be *continually increasing*. They can start with measures to reduce the spread of the disease, such as social distancing and facemasks in the case of Covid-19, and expand to increasing numbers of hospital beds as needs continue, and to methods to help people recover, but must continue to vaccines to further reduce the spread, possibly to drugs to reduce susceptibility or infectivity, and to continual quests for antimicrobials to cure those infected. Relaxing on the pursuit of new efforts of control may not result in the green line in Figure 6.

## VII. Discussion

The *rsN* method has at least two major benefits. Exponential population growth is often linearized by plotting it on a log scale. However logarithmic lines may not be broadly intuitive and they do not offer a glimpse of mechanism, as per-capita growth rate does. Second, modelers know that exponential growth is never possible over the long-term [7]; the curve will eventually level out. We should ask why a basic formulation like the *rsN* model should work. As a model is refined—with more parameters, deeper mechanisms, and greater spatial resolution—at least three uncertainties come into play.

### A. Model errors

There may be uncertainties in the deeper mechanisms that are added to the more complex model, so that the mechanisms do not precisely mimic the dynamics of the actual system being modeled. For example, the process might be modeled as either density-dependent or frequency-dependent transmission, when in fact the actual transmission may be a blend of those two.

### B. Data errors

Data required to parameterize a more complex model may not be available or accurate. For example, as mentioned, the actual prevalence in a model of a disease with an asymptomatic phase cannot be known unless an assay for the infection is widely available. With more and more parameters in a model, statistical variance in estimates of the parameters can add up to overwhelm the model. This is related to the statistical property that the variance of a sum is the sum of all the individual variances.

### C. Sampling errors

Finer resolution in a model will reduce the number of data points observed for each category, eventually to a level where inevitable statistical errors in sampling can combine with nonlinearities in the model to make an estimation of the parameters intractable and to thereby produce erroneous results.

The first two of these uncertainties involve science in constructing the model and determining its correctness, and in understanding where the data being gathered may not completely represent the process being modeled. The third of these is purely statistical, given a model and its parameters. For any specific case, it is possible to use statistical partitioning into multiple simulations to see how subdivisions of the model, for example into smaller and smaller geographic regions, might introduce errors in more complex models that are not inherent in simpler ones.

Finally, the projections of the number of US deaths from Covid-19 we present here are considerably smaller than some other estimates that have been projected. However, that must not be taken as a reason to relax efforts to subdue the spread. On the contrary, it is evidence that the expanding measures are working, and they are visible as a negative *s* term in the model. Relaxing the measures as deaths increase, or as time passes, before the disease is subdued, will change the value of the *s* term and increase the projections offered here. The cumulative number of deaths can approach a limit, *− r*/*s*, when measures continue to expand. If measures don’t continue in this way, these projections from real data will not hold. Continuing future applications of this model using deaths can reflect, with some delay, when measures represented by *s* are not keeping pace with what is needed.

## Data Availability

All data are from publicly available sources.

We thank Peter White and David Tilman for encouraging an early draft of this paper, Adrienne Keen for providing comments on the Ebola section, and Todd Lehman for applying his data-smoothing software to Ebola data in Figure 1B.

## Appendix I Cumulative deaths as a dynamical variable

Using the cumulative number of deaths as a dynamical variable may seem unwarranted because past deaths do not feed forward to cause new infections. This appendix explains why this actually is warranted, and doing so is useful because deaths are one of the few variables that can be estimated with a degree of accuracy early in an outbreak.

**Equation 1**. The starting point is an *SIR* epidemiological model, augmented here to also track deaths. *S* = *S*(*t*) is the number of individuals in the population at time *t* who are susceptible to the disease, *I* = *I*(*t*) is the number infected, *R* = *R*(*t*) the number once infected but now recovered, and *N* = *N*(*t*) the number who have died. Both *R* and *N* cumulative values, with flows in but not out of the categories. Parameter *β* is the infectivity, *γ* the rate of recovery, and *α* the rate of death from the disease. The values of most of those parameters and variables cannot be known accurately at the beginning of an outbreak. Functions *g*_*i*_(*t*) represent other population processes such as births, deaths, or migration.

**Equation 2**. The central part reduces to an *I* model. Suppose that a new outbreak begins with a single infection and spreads from low levels based on its transmission parameters. Suppose also that most of a large population is susceptible, so that the term *S*/(*S*+*I*+*R*) is essentially 1. Further suppose that the disease spreads rapidly relative to ordinary population processes such as births, so that population changes embedded in functions *g*_*i*_(*t*) will be negligible. In that case the *SIR* model reduces to an *I* model that is approximately dependent only on the infectivity *β*, the recovery rate *γ*, and the death rate *α*. This reduced equation has an explicit solution *I*(*t*), which appears to the right of the arrow. The multiplicative constant of integration is unity because the number of infections at *t* = 0 is 1.

**Equation 3**. The total number of deaths, *N*, accumulates in proportion to the number of infections. The number of infections has an explicit solution and the differential equation for the number of deaths has an explicit solution too. That solution appears to the right of the arrow. The term *−* ‘1’ is a constant of integration arising because the number of deaths at *t* = 0 is 0.

**Equation 4**. The relative rate of change in infections, *dI*/*dt* (left part of Equation 2) divided by *I* (right part of Equation 2, integrated), which is *r* = *β − γ − α*.

**Equation 5**. The relative rate of change in cumulative deaths is *dN*/*dt* (left part of Equation 3) divided by *N* (right part of Equation 3, integrated), which is approximately *r* = *β − γ − α*.

In all of the equations above, adjusting *β* downward represents efforts to control the spread of the disease, such as isolation and face masks in the present pandemic. Adjusting *α* downward represents better care to reduce mortality, for example increasing the abundance of respirators in hospitals. Adjusting *γ* upward also represents better care, helping those infected recover more quickly. It can also represent antibiotics that can cure individual already infected. However, vaccination is not provided for in *SIR* Equation 1. That would be handled by an additional term *− κS* in *dS*/*dt* and +*κS* in *dR*/*dt*, directly connecting the susceptible *S* category to the *R* category—*R* then meaning resistant rather than recovered.

In summary, because cumulative deaths *N* grow in close proportion to the number of present infections *I*, we may construct a dynamical system based on cumulative deaths that represents the dynamics of the *SIR* system when the proportion of susceptible is high, as in the early stages of an outbreak. It can be shown by simulation that this extends beyond the early phases and to more general conditions, as illustrated in this paper.

## Appendix II Sub- and super-exponential growth

The observed relative rate of spread across a region or around the globe can oscillate, sometimes apparently spreading faster than exponentially and appearing to be out of control, while at other times spreading slower than exponentially and appearing to be moderating. Part of this can be due to new regions becoming infected that differ in their degrees of unpreparedness for such an event. Another part, however, can simply be intrinsic to the dynamics of a spreading pandemic. This appendix illustrates numerically how the latter can arise. It is related to the general topic of ecological metapopulation models [16].

Suppose that when an infection reaches a new region, it starts to spread through the region at a decreasing rate, because of increasing care in local neighborhood interactions and other means, as happens in Equations 2 and 3 with a negative *s*. Further suppose that sometime during its spread through a region, some infected members carry it to different regions, through long-distance interaction such as travel by air or rail.

The data table below starts with a single infected region, column C01, that grows logistically from a single infection ultimately to 1000 infections, but about halfway through the expansion, it seeds two other uninfected regions, columns C02 and C03, within which the infection also starts spreading logistically. Later the three now-infected regions each seed one uninfected region on average, now making six total infected regions. Later still the six now-infected regions each seed less than one new region each. Thus the spread between regions is slowing, from 2 new regions for each infected region to 1 new region to about 0.8, and the rate of spread within each region is also continually moderating following logistic dynamics. Nonetheless, across all regions combined, the disease can grow faster than exponentially at times, as it is subsiding (Figure 10).

**Figure 9:**
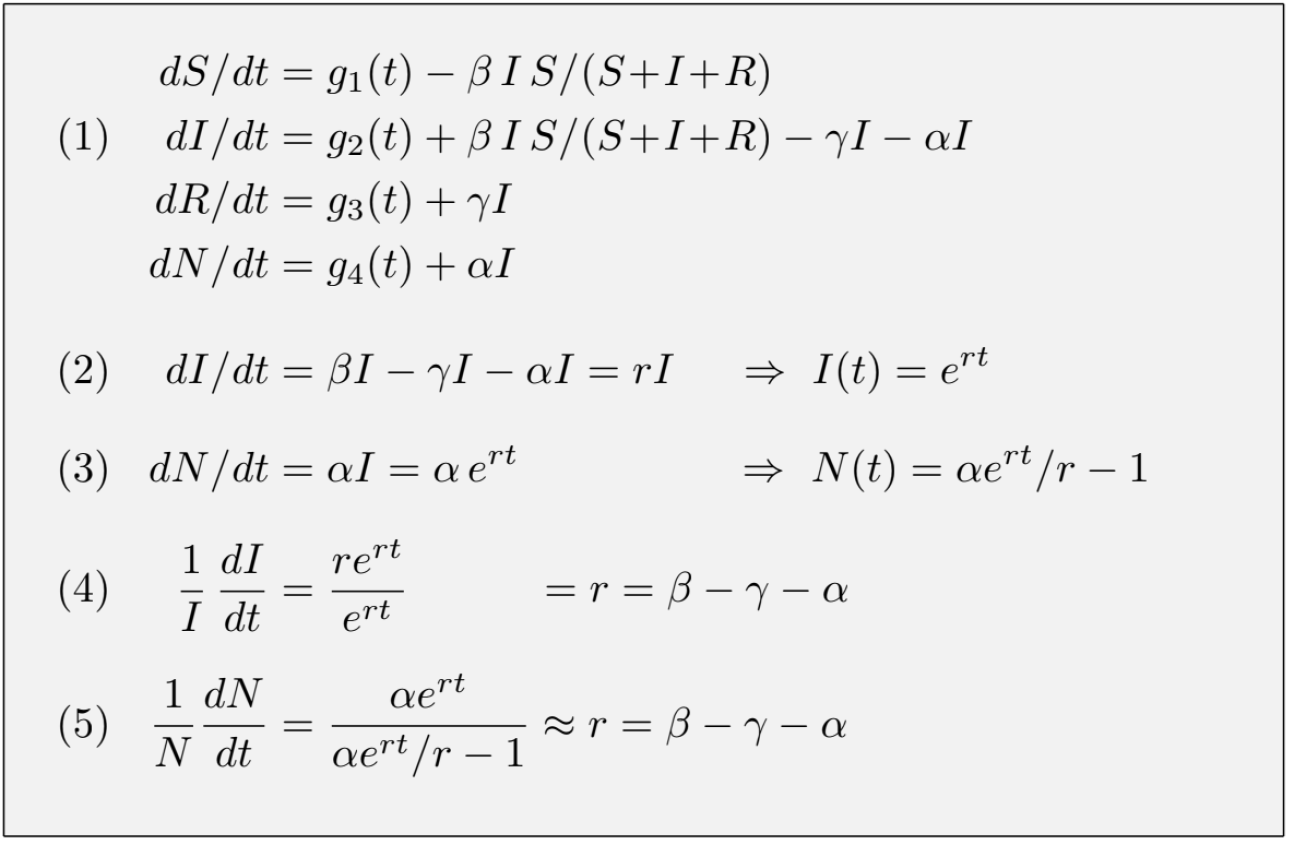
Proportionality of infections and cumulative deaths

**Figure 10:**
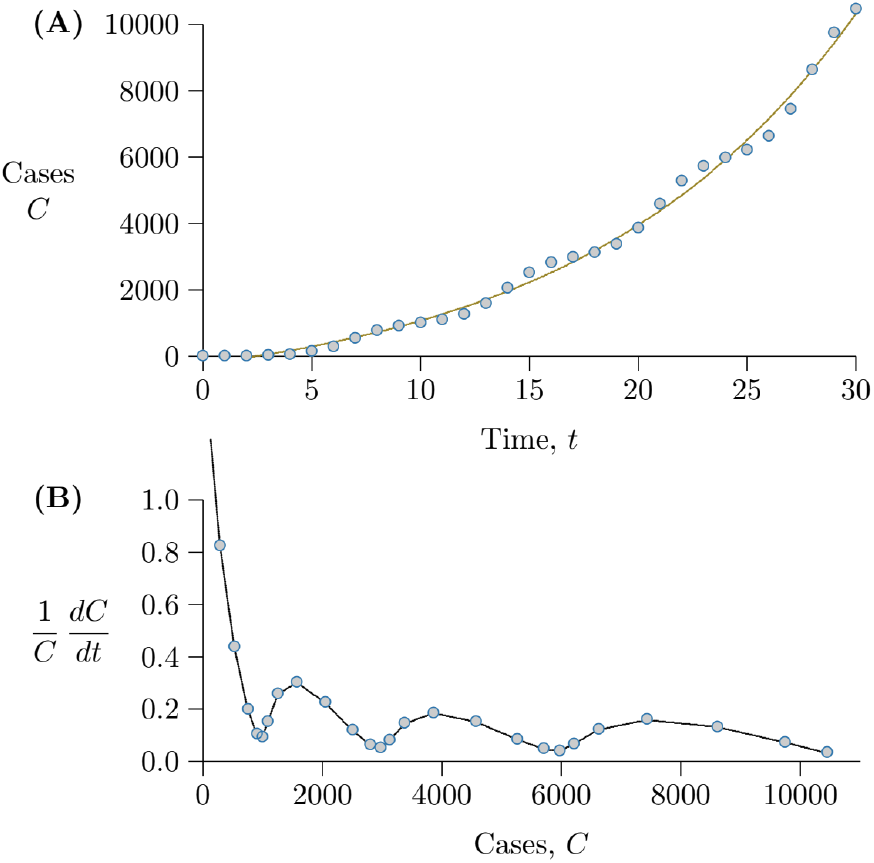
Local growth and regional spread graphically. (A) From the right-most column of 11, representing all regions combined, the expansion appears to be exponential. It is not, even though an exponential curve through the data fits with R^2^ > 0.99, (B) The same data graphed as a per capita growth rate, revealing the curve in Part A as sub-exponential, though with a signature of repeated blips of super-exponential and sub-exponential growth. (See Figures 7 and 1B for similar signatures in actual data from Ebola (2014–15) and Covid-19).

**Figure 11:**
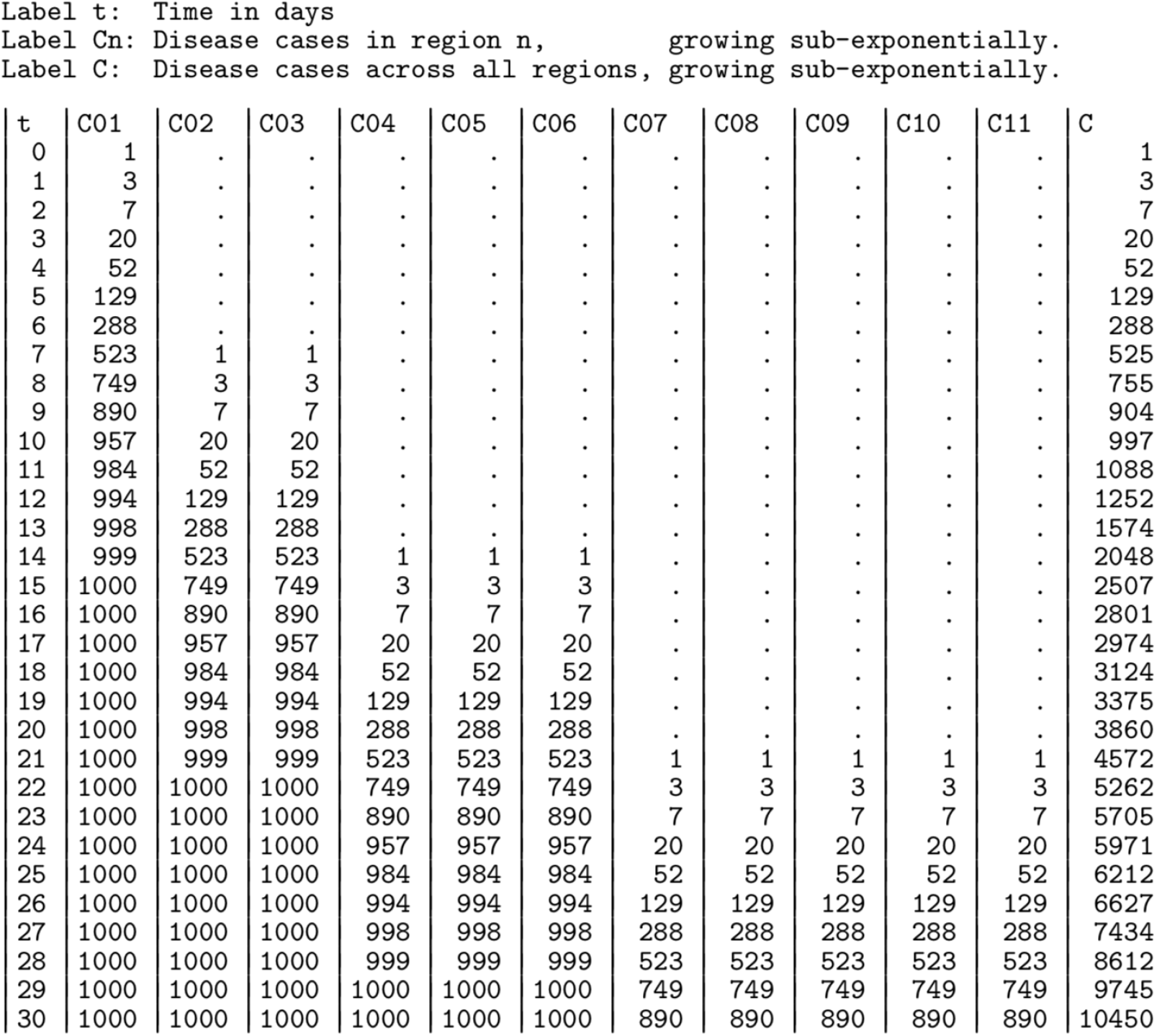
Local growth and regional spread of a fictional disease.

**Figure 12:**
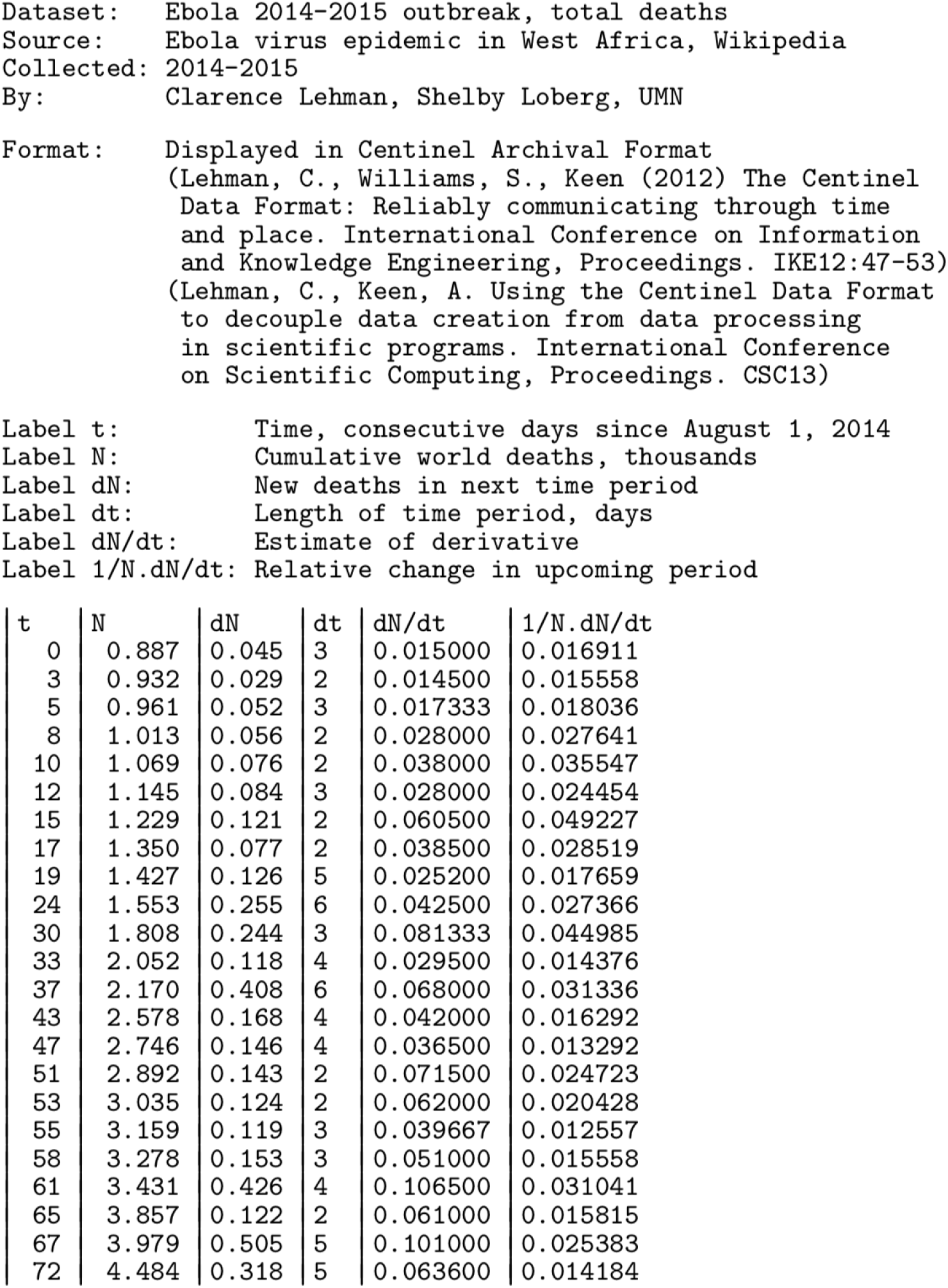
Early data for Ebola (2014–15)

## Appendix III Early data for ebola (2014–15)

## Notes

### Competing Interest Statement

The authors have declared no competing interest.

### Funding Statement

Work was supported in part by the University of Minnesota Institute on the Environment. No external funding was received.

